# Prevalence, clinical correlates and outcomes of cardiorenal anemia syndrome among patients with heart failure attending tertiary referral hospital in Dodoma, Tanzania: A protocol of a prospective observational study

**DOI:** 10.1101/2023.09.28.23296304

**Authors:** Gidion Edwin, Baraka Alphonce, Alfred Meremo, John Meda

## Abstract

**Background:** Cardiorenal anemia syndrome (CRAS) is a common complication among patients with heart failure and is associated with poor clinical outcomes. However, there is a paucity of published data concerning CRAS, despite of significant increase in heart failure patients attending medical services in developing countries. This study aims to assess the prevalence, clinical correlates, and outcomes of CRAS among patients with heart failure attending the Benjamin Mkapa Hospital in Dodoma, Tanzania.

**Methodology:** A prospective observational study is ongoing at the Benjamin Mkapa Hospital in Dodoma, Tanzania. Currently, the study has enrolled 68 patients. The socio-demographic data, clinical correlates, and prevalence of CRAS will be determined at baseline meanwhile, the outcomes of CRAS will be determined during a follow-up period of six months from the date of enrollment. CRAS is the primary outcome of the study. Data will be categorized into CRAS and non-CRAS during analysis. Mean and standard deviation will be used for normally distributed continuous variables while median and interquartile range will be used for skewed data. Frequencies and percentages will summarize categorical variables. Clinical correlates and outcomes of CRAS will be analyzed and compared by using univariate and multivariate logistic regression models. A two-tailed p-value of less than 0.05 will indicate statistical significance.

## Introduction

Cardiorenal anemia syndrome (CRAS) is public health concern worldwide among heart failure patients and is associated with poor clinical outcomes [1,2]. The pathophysiology of cardiorenal anemia syndrome is complex and not clear [2–4]. The reported prevalence of CRAS among heart failure with reduced ejection fraction ranges 4.6% and 35.4% in developed countries [5–7] and 19% to 66% in developing countries [2,5,8].

CRAS management poses great challenging due to increased cost of services and disease-drug interactions, which portends poor prognosis [1,2,9] which explains a very high CRAS related mortality in developing countries (73.5%) than in developed countries (9.5%) [1,2,10]. There are several clinical correlates which have been reported to contribute the development and worsening of CRAS including among others diabetes, hypertension, dyslipidemia, obesity, alcoholism and iron deficiency with or without anemia but all these correlates have not yet established in our settings [5,11–13]. The most common outcomes of CRAS includes cardiovascular death, poor quality of life, worsening heart failure, worsening anemia and worsening renal function [1,2,14].

There is a significantly increasing burden of HF patients in Tanzania. However, there is unpublished data from hospital registries pertaining cardiorenal anemia syndrome [5,9,15]. This study is the first to be conducted in our settings and will determine the prevalence, clinical correlates and outcomes of cardiorenal anemia syndrome among the patients with heart failure irrespective of ejection fraction attending the Benjamin Mkapa hospital in Dodoma, Tanzania.

## Materials and methods

### Study objectives

1. To determine the prevalence of cardiorenal anemia syndrome among patients with heart failure attending a tertiary referral hospital in Dodoma, Tanzania
2. To determine the clinical correlates of cardiorenal anemia syndrome among patients with heart failure attending a tertiary hospital in Dodoma, Tanzania
3. To determine the outcomes of cardiorenal anemia syndrome among patients with heart failure attending a tertiary referral hospital in Dodoma, Tanzania

### Study design

A hospital based prospective observational study

### Study duration

Study is conducted for 9 months period start from 18^th^ August, 2023 to 18^th^ April, 2024. Recruitment is from 18^th^ August, 2023 to 18^th^ October, 2023 and follow-up period will commence a month later after recruitment up to 6 months. Last patient will followed up to 18^th^ April, 2024.

### Study settings

This study is ongoing at the Benjamin Mkapa Hospital (BMH). This is a government tertiary hospital that serves as a national consultant hospital located in the capital city of Tanzania. The hospital is located within the University of Dodoma (UDOM) premises, 13 kilometres from Dodoma city center. It was established in 2015 and named in remembrance of Former third President, His Excellence Benjamin William Mkapa. This hospital serves a population of Dodoma of about 3.08 million as per Tanzania demographic health survey (TDHS) report published in 2022 [16] and other inhabitants from neighbouring regions of Iringa, Singida, Manyara, Tabora, Morogoro,, and Katavi [17]. The hospital has bed capacity of 400. The BMH serves as a teaching hospital for the University of Dodoma for both undergraduate and graduate programs.

### Study population

Patients (Human subjects) with heart failure who met inclusion criteria will be recruited.

### Inclusion criteria

1. Patients aged ≥ 18 years with a diagnosis of Heart failure irrespective of ejection fraction with echocardiographic report within 6-months prior enrollment.
2. Patients with chronic heart failure attending respective clinics
3. Patients admitted with a confirmed index diagnosis of acute heart failure by Framingham criteria and cardiac imaging
4. Patients with acute on chronic heart failure
5. Patients with either baseline evidence of kidney disease based on elevated serum creatinine within 6-months from prior enrollment or new patient with Chronic kidney patient
6. Patient who has signed the informed consent form or proxy consent incase the patient is incapable

### Exclusion criteria

1. Patients with known solid or hematological malignancy on chemotherapy or radiotherapy
2. Pregnant women
3. Patients with obvious gastrointestinal bleeding from any cause
4. Patients with established Hemoglobinopathies or other non-gastrointestinal bleeding disorders
5. Patients with known history of Chronic kidney disease stage 5 on iron supplementation, or on renal replacement therapy
6. Patients underwent implantable cardiac devices or pacemaker implantation

### Sample size estimation

To estimate the sample size, the proportionate formula used in prospective cohort studies will be used [18]

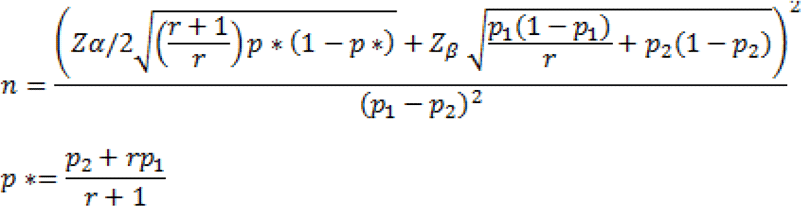

Where by

r = ratio between the two groups

p1 = CRAS prevalence obtained from literature

p2 = expected prevalence of CRAS from the study

p1 –p2 = effect size

Zβ = standard normal variate for statistical power

Zα/2 = standard normal variate for significance level

The prevalence of CRAS in previous studies was 44.4% [2]

The prevalence of CRAS in this study is expected to be 48% Therefore;

r = 1

p1 = 44.4%

p2 = 48%

p1 – p2 = 18%

Zβ = 0.84 for statistical power of 80%

Zα/2 = 1.96 for significance level of 95%

n= 119

Considering the 30% attrition rate adopted from previous study [2]

Therefore, the minimum sample size required to meet the objective was 160 patients.

### Clinical outcomes

#### Primary outcome

Cardiorenal anemia syndrome (CRAS) is the primary outcome of this study and will be defined as a triad comprising heart failure regardless of ejection fraction, chronic kidney disease with estimated glomerular filtration rate <60mil/min/1.73m^2^ and anemia of <12 g/dl for women or <13 g/dl for men [1,2]

#### Secondary outcomes

Secondary outcomes will include:

##### Cardiovascular death

is defined as any cardiovascular related death occur within follow –up period [2,19,20].

##### Worsening heart failure

is defined as deterioration in functional class as identified by New York Heart Association (NYHA) criteria or Heart failure hospitalization rate more than one within follow-up period or change in ejection fraction to lower scale than baseline by echocardiography or both [2,3,5].

##### Worsening renal function

is defined when there is a change in chronic kidney disease stage to lower stage than baseline or a need of a dialysis or both [21].

##### Worsening anemia

is defined when there is a change of hemoglobin level to lower level from baseline which will then need either blood transfusion or erythropoietin use or both [3,22].

### Study variables

#### Objective 1

Aims to determine the prevalence of cardiorenal anemia syndrome among patients with heart failure irrespective of ejection fraction at baseline.

**Table 1:**
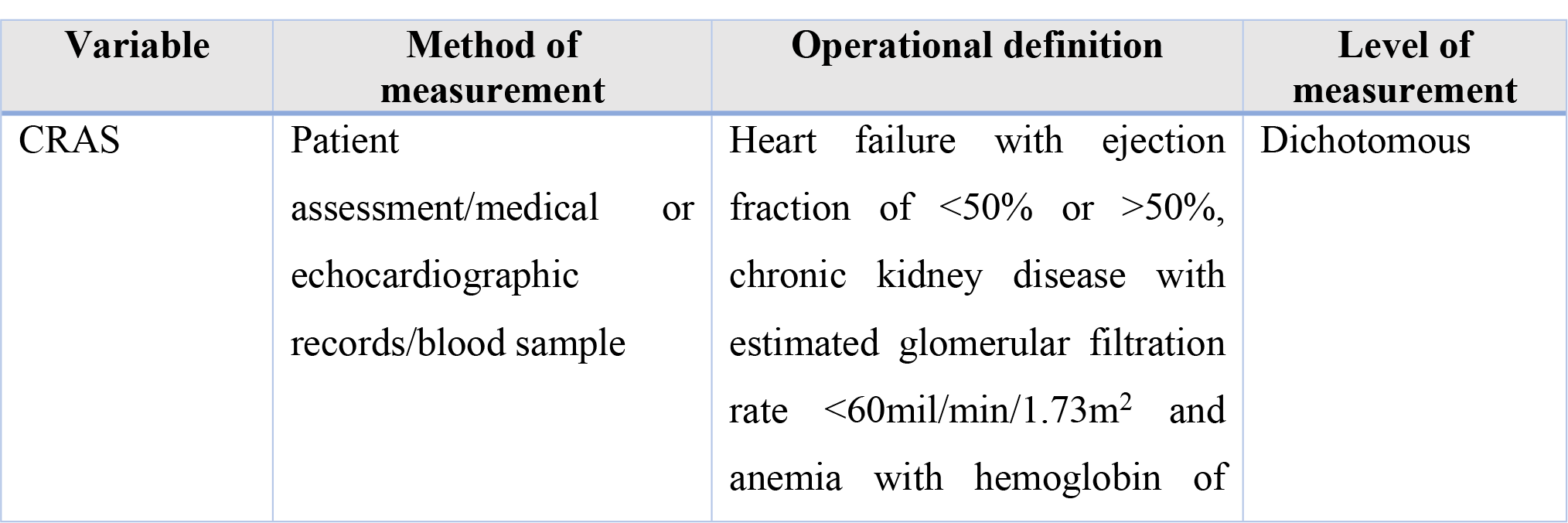
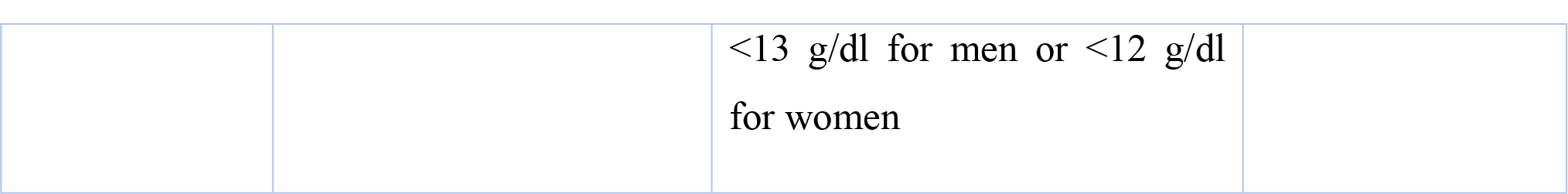
Summary and description of variables for the objective number 1.

#### Objective 2

Aims to determine the clinical correlates for the cardiorenal anemia syndrome at baseline among the patients with heart failure irrespective of ejection fraction [2,7,12]. Information from medical records such as chronic kidney disease using serum creatinine level and echocardiographic report will be accessed from 18^th^ February, 2023 onwards.

**Table 2:**
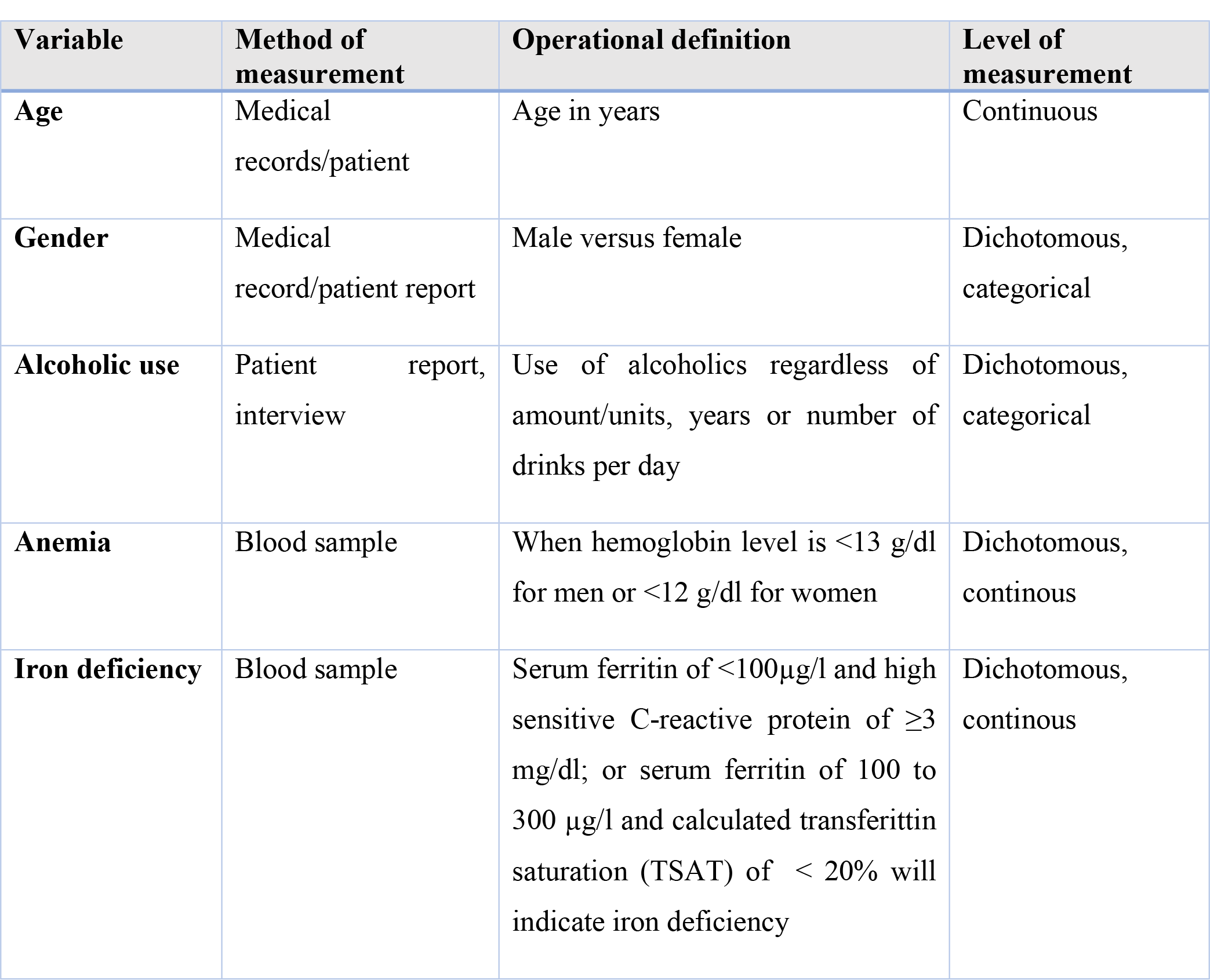
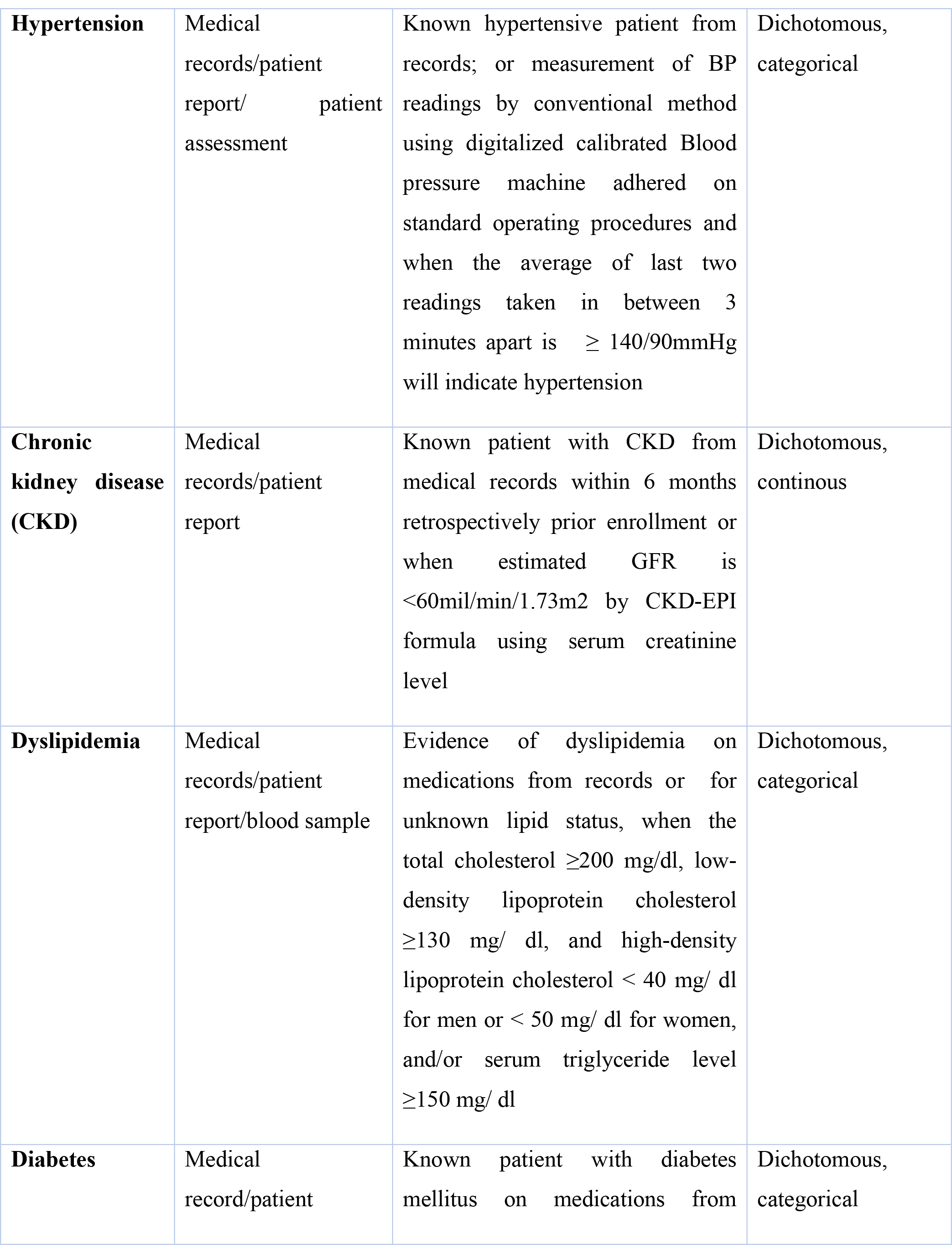
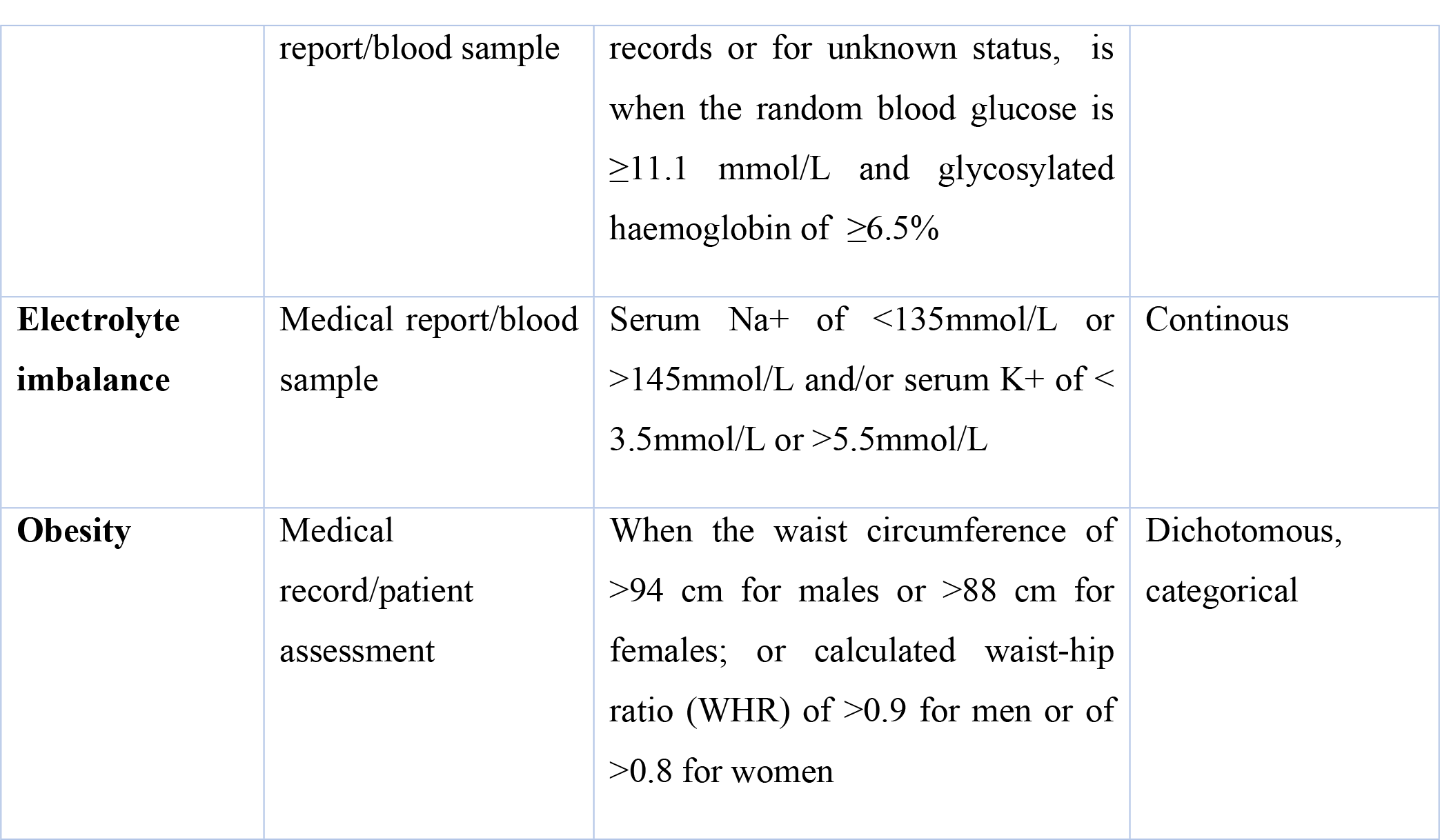
Description of the clinical correlates.

#### Objective 3

Aims to determine the outcomes of the cardiorenal anemia syndrome among the patients with heart failure within six months follow up period [2,5,7,12,26].

**Table 3:**
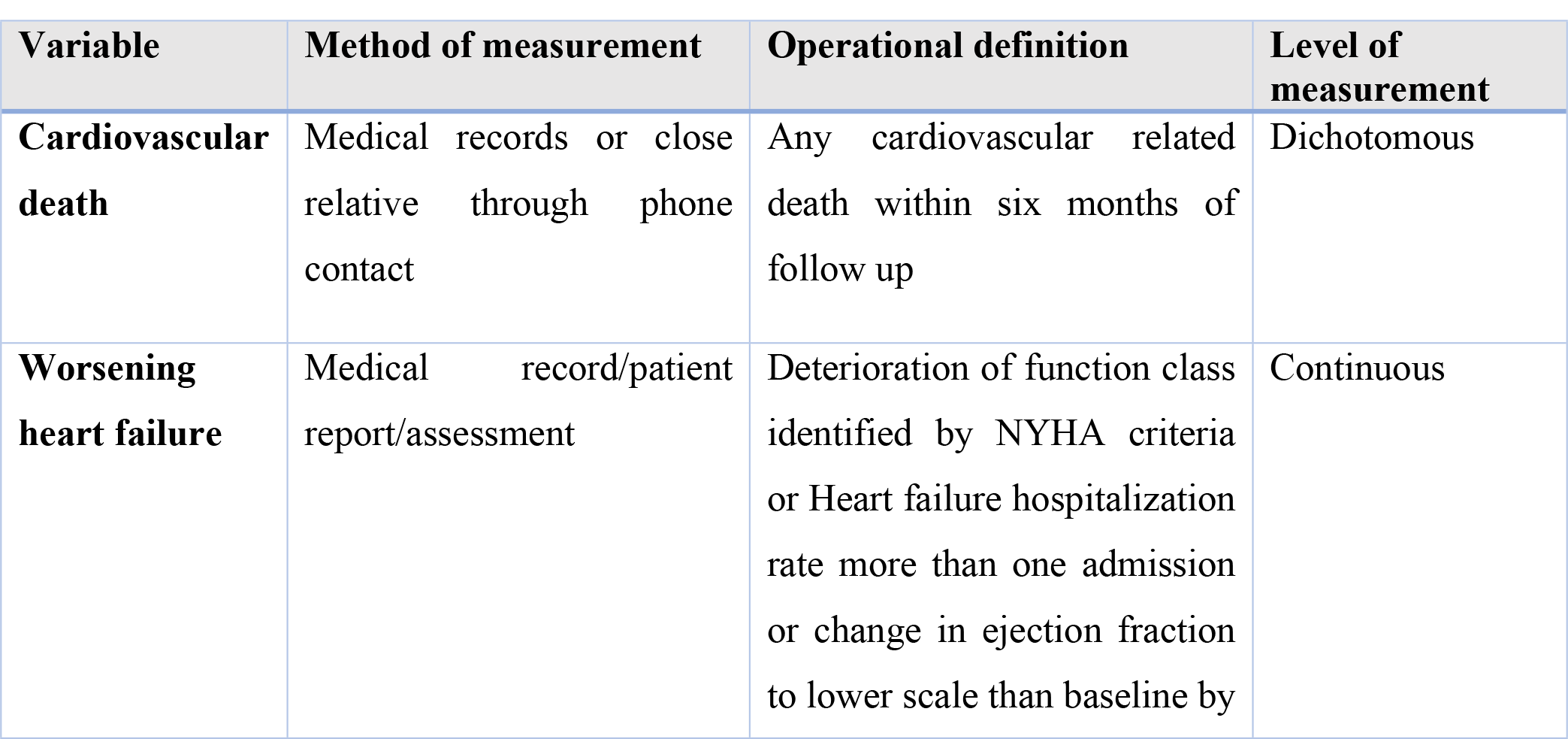
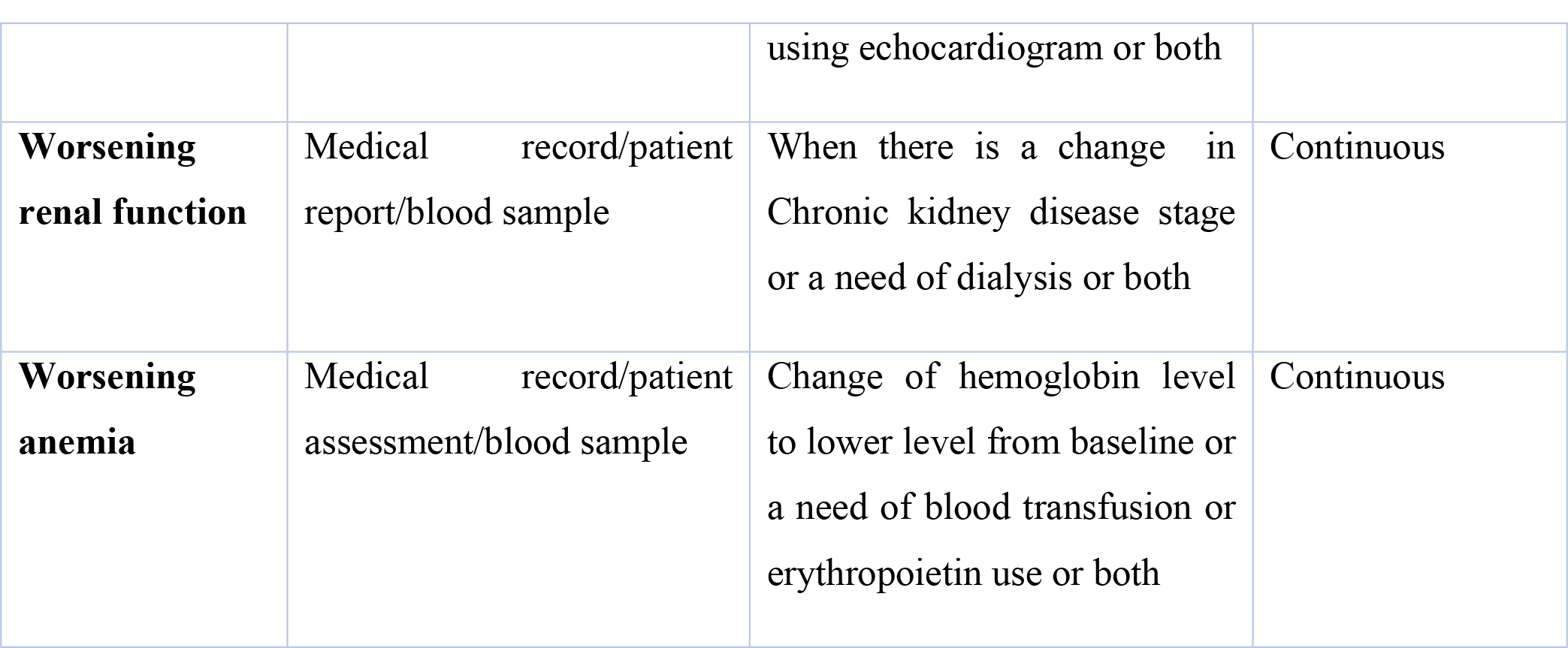
Description for the clinical outcomes.

#### Participants’ characteristics

Participants are adults of 18 years or older attending internal medicine, cardiology and nephrology departments of the Benjamin Mkapa hospital in Dodoma who met the study criteria [4,19,27].

### Data collection process

#### Evaluation of the participants

A minimum of 160 patients who meet the inclusion criteria will be enrolled in this study from 18^th^ August, 2023 to 18^th^ April, 2024 including 3-months of enrollment and a 6-month of follow up. Follow-up period will be done on monthly basis via phone calls or through routine clinic visits to assess for the secondary outcomes. Data will be collected using a well-structured questionnaire, which will include socio-demographic information, physical examination and anthropometry, assessment of underlying comorbid conditions, laboratory investigations, and non-invasive cardiac imaging. Socio-demographic data will include age, gender, smoking history, history of alcoholic use either current or previous case, and anthropometric measurements including waist circumference, hip circumference and then, waist-hip ratio will be calculated [28]. Focused clinical history will be exhausted to assess the symptoms related to heart failure using modified Framingham criteria. Two major criteria or one major and two minor criteria will be sufficient to make a diagnosis of heart failure [29]. Subjective Functional status by using New York Heart Association functional criteria will be recorded, which comprises four classes as follows [2,13,30]: class 1 means no limitation to physical activities, class 2 means slight limitation to strenuous activities, class 3 means marked limitation to simple and slightly activities and class 4 means symptoms at rest.

#### Measurements of clinical variables

##### Waist circumference

is the measurement of the abdominal girth in centimeters and will be measured in erect position with abdomen relaxed and arms aside and will be recorded over the unclothed relaxed abdomen at the smallest diameter between the coastal margin and the iliac crest of which >94 cm for men and >88 cm for women will be defined as central obesity. In addition, hip circumference in centimeter will be recorded over minimal clothing at the level of the greater trochanter (usually the widest diameter around the buttocks). Waist-hip ratio (WHR) is dimensionless measurement and will be calculated from waist circumference and hip circumference. A WHR ≥0.9 or ≥0.8 will indicate obesity. The sensitivity and specificity of waist-hip ratio of more than 85% compared to body mass index (BMI) in predicting obesity [28,31].

##### Blood pressure measurement

a blood pressure (BP) reading will be measured by the conventional method by using a calibrated automated BP machine keeping with the 2018 AHA/ACC hypertension guideline for standard measurement of BP. The patient should be required to empty the urinary bladder prior to measurement, not to smoke, not take alcoholics or caffeine nor strenuous exercises within 30 minutes prior to measurement. Three BP readings will be taken in relaxed position 3 to 5 minutes apart per each reading and an average of the last two BP readings will be used. Blood pressure ≥ 140/90 mmHg will be defined as hypertension [2,17,32].

#### Laboratory and imaging investigations

##### Collection of blood sample

The laboratory standard operating procedures accredited and supervised by the Benjamin Mkapa hospital will be warranted throughout the study. Each participant will consent for venipuncture which will inflict pain during sample collection, then laboratory technician will follow the aseptic procedures during sample collection. Before sample collection, collecting tubes (purple toped EDTA (K2/ K3) sodium fluoride plain with additive and another red toped plain tubes, with no additive) will be labeled with the patient’s particulars. The tourniquet will be applied 30 to 60 centimeters above elbow joint. The intended site of venipuncture will be cleaned with 70% methylated spirit and the area is left to dry for 15 to 20 seconds before venipuncture. Then 10 mls of venous blood will be taken using a 10-cc syringe.

The tourniquet will be released and removed, and the site of venipuncture will be compressed with a cotton swab for 1 to 5 minutes to arrest bleeding. From the samples, the following tests will be analyzed: lipid profile (high density lipoprotein-cholesterol, low density lipoprotein-cholesterol, total cholesterol and serum triglycerides), full blood picture (haemoglobin level, mean corpuscular volume, mean corpuscular hemoglobin), Iron studies (serum iron, serum ferritin, total iron binding capacity and transferrin saturation), high-sensitive C-reactive protein, and serum creatinine and serum electrolytes (serum sodium and serum potassium). The cool box will be used for transportation of samples to the BMH laboratory for processing 15 to 30 minutes after sample collection. The serum sample will be separated from whole blood by centrifugation at 300 rpm for 5 minutes, and two aliquots will be prepared. A sample is stored at 2°c– 8°c if the analysis is expected after 2 hours from sample collection especially for iron studies which be analyzed after every 100 collected samples using MAGLUMI 800 CLIA. However, blood samples will be stored at room temperature if the analysis is expected to be done within 2 hours from time of sample collection. Analysis of the sample will be done using clinical chemistry automatic analyzer MAGLUMI 800 CLIA made in China, 2019 and Roche Cobas 6000 CLIA made in USA, 2018.

##### Lipid profile

Dyslipidemia is defined as total serum cholesterol ≥200 mg/dL, or LDL-cholesterol ≥130 mg/dL, or serum triglyceride ≥150 mg/dL, or HDL-cholesterol <40 mg/dL for women and <50 mg/dL for men [17,33]

##### Blood Sugar measurement

Diabetes mellitus will be defined as either known patient with type 1 or 2; and for unknown diabetic status, is when random blood glucose by using blood from finger prick is ≥11.1mmol/L and glycated hemoglobin is ≥6.5% [15,34]

##### Iron studies

Iron deficiency will be defined as serum ferritin of <100μg/l and high sensitive C-reactive protein of ≥ 3 mg/dl [35], or serum ferritin of 100 to 300 μg/l and transferittin saturation (TSAT) ≤ 20% [7,11–13,36].

##### Full blood picture

anemia will be defined when hemoglobin ≤ 12g/dl for women or ≤ 13g/dl for men. Mean Corpuscular Volume (MCV) and Mean Corpuscular Hemoglobin (MCH) will be recorded to characterize the type of anemia [2,17,37].

##### Serum electrolytes

electrolyte imbalance will be considered when the serum sodium is ≤ 135 mmol/l (hyponatremia) or ≥ 145mmol/l (hypernatremia) and / or potassium level of ≤ 3.5mmol (hypokalemia) or ≥ 5.3 mmol/l (hyperkalemia) [21,25,33].

##### Blood creatinine

Chronic kidney disease will be defined following estimation of glomerular filtration rate. Glomerular Filtration Rate will be calculated by using CKD-EPI formula by adjustment of African-American correction factor and value of <60mil/min/1.73m2 will be considered as chronic kidney disease [25,34,38].

#### Two-dimension transthoracic echocardiogram (2D-TTE)

Transthoracic Echocardiography (model Vivid TM T9 manufactured by GE Healthcare, USA 2018) will be used for recommended patients who are either newly diagnosed patients or for previous known HF patients of which their echocardiogram reports are of more than 6-months prior to index contact. Transthoracic Echocardiogram will be done by experienced echocardiographer and/or confirmed by consultant cardiologists. Depending on condition of the patient so, a patient will be instructed to undress and expose the chest then, the patient will lie on the left lateral decubitus position and then, change to supine position during imaging for different echocardiogram views. The imaging will take 10 to 20 minutes.

During imaging, the confidentiality and privacy will be warranted. Findings of potential diagnostic importance will be extracted from the echocardiogram report which will include diastolic dysfunction of either grade, elevated left atrial pressure and left ventricular ejection fraction [27]

#### Follow-up procedure

Every patient will be followed up on monthly basis within 6-months via phone calls or during regular clinic visit. In case a participant is not reachable following 3 phone calls on 3 alternative days on every fourth week of follow-up month or during clinic visit. Failure to be reacheable will indicate lost to follow-up.

### Data management

All collected data will be accessible to research team only. All completed questionnaires will be coded before entered into spreadsheets, and patients will be identified by unique patient’ registration number of the BMH linked to patient name, age and sex which will be stored in encrypted secured computer,

### Safety consideration

Throughout this study, the safety of the patients will be highly observed. Only minimal injury and pain is expected during venipuncture. During echocardiogram, the patient needs to undress the chest for procedure so, privacy will be ensured. All patients will receive high quality of care as per national and hospital guidelines despite of his or her readiness to participate in this study. No minors will be enrolled.

### Data analysis

All completed questionnaires will be coded before entered into dataset on Microsoft excel. SPSS windows version 25 program (IBM SPSS, Chicago IL) will be used for data management and analysis. Data will be classified as CRAS or non-CRAS groups. Data will be presented as means **±** standard deviations for continuous variables with normal distribution, or median and interquartile range for no-parametric (skewed) variables [1,2,14]. Categorical variables will be presented as frequencies and percentages. Socio-demographic variables and clinical correlates of cardiorenal anemia syndrome at baseline will be analyzed and compared using Pearson chi-squared test for the categorical variables, and independent student’s t-test for the continous variables. [1,2,10]. The clinical correlates including age, gender, alcohol use, smoking history, hypertension, obesity, diabetes, and dyslipidemia and iron deficiency will be analysed and compared by using univariate and multivariate logistic regression. Cox proportional hazards model will be used to analyze and compare the outcomes of CRAS. A two-tailed p - value of less than 0.05 will indicate statistical significance [2,7,14].

### Ethical issues

The study complies with ethical principles that have their origin in the Declaration of Helsinki. The ethical clearance to conduct the study is obtained from the institutional research review ethical committee (IRREC) under the Vice chancellor’s office of the University of Dodoma given a reference number (MA.84/261/64/123). The approval and permission for data collection is obtained from the Benjamin Mkapa hospital.

### Study Timeline

Study duration is 9-months commencing from 18^th^ August, 2023 and will end on 18^th^ April, 2024.

## Discussion

Globally and in Tanzania, there is significantly step-rise burden of heart failure patients attending medical services and most of these patients succumbs CRAS which then, portends poor clinical outcomes [2,39]. Reported data of CRAS irrespective of ejection fraction is significantly higher in developing countries as compared to developed countries [1,2,39]. In our settings, there is paucity of data concerning CRAS despite of having many patients with heart failure attending medical services. The ongoing prospective observational study is assessing the prevalence, clinical correlates and outcomes of CRAS. Establishment of the burden of CRAS, clinical correlates and outcomes in our settings will be a benchmark towards the large studies with high level of evidence and recommendation in order to improve the health care delivery systems so as to reduce the poor clinical outcomes and hence, improves quality of life.

## Data Availability

No datasets were generated or analysed during the current study. All relevant data from this study will be made available upon study completion.

## Acknowledgments

The authors will appreciate the patients and staffs of the Benjamin Mkapa Hospital in Dodoma, Tanzania for their readiness and voluntarily active participation in this study.

## Author contributions

**Conceptualization:** Gidion Edwin, Alfred Meremo, John Meda

**Data clearance and curation**: Gidion Edwin

**Formal analysis**: Gidion Edwin, Baraka Alphonce

**Investigation:** Alfred Meremo, John Meda

**Methodology**: Gidion Edwin, Baraka Alphonce, Alfred Meremo, John Meda

**Supervision**: Baraka Alphonce, Alfred Meremo, John Meda

**Writing – original draft**: Gidion Edwin

**Writing – review & editing:** Baraka Alphonce, Alfred Meremo, John Meda

---

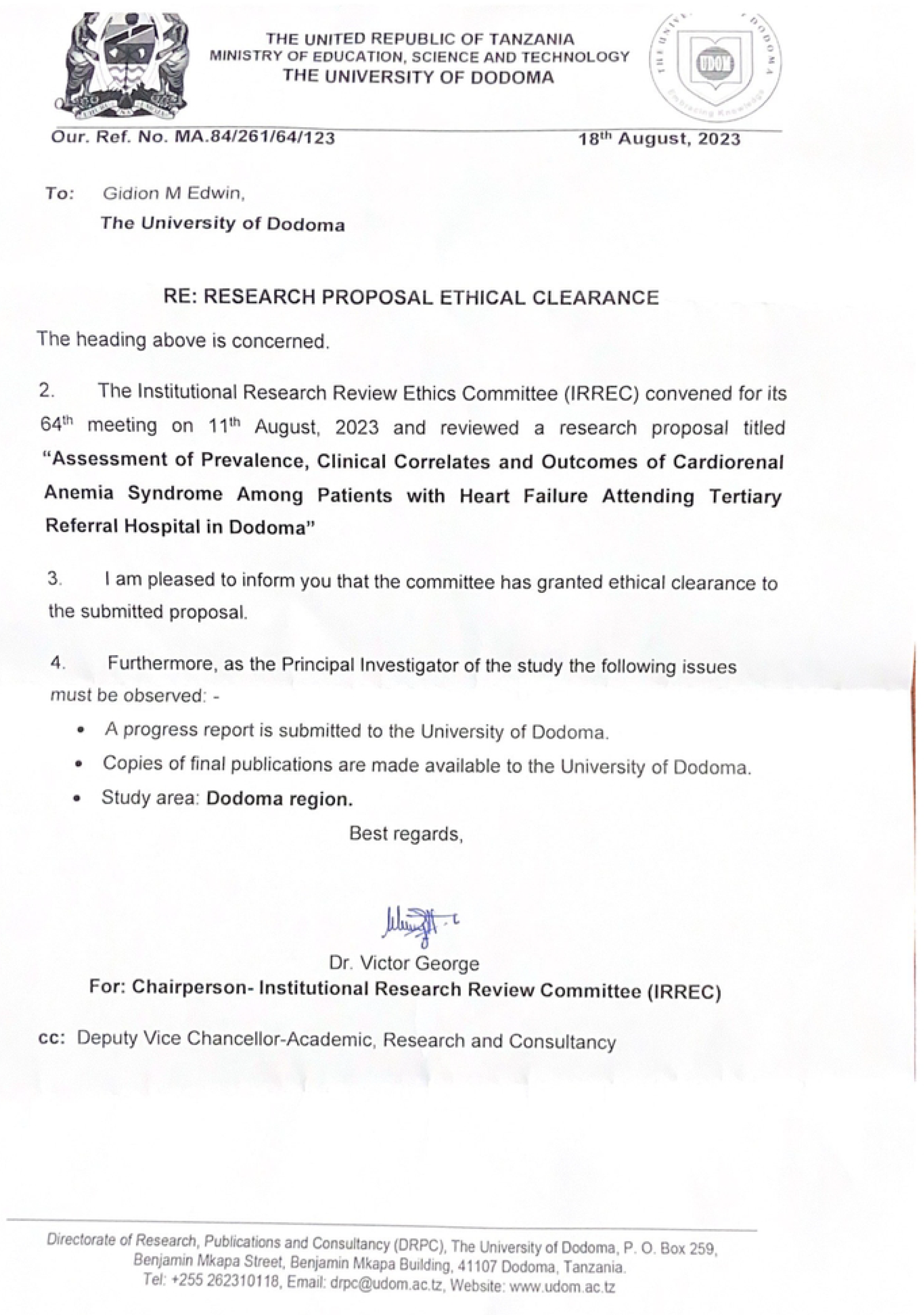

